# Dissatisfaction with family members’ medical care: the relationship with trust in personal physicians and physicians generally in Japan

**DOI:** 10.1101/2021.04.21.21255773

**Authors:** Nao Oguro, Ryo Suzuki, Nobuyuki Yajima, Kosuke Sakurai, Takafumi Wakita, Mark A. Hall, Noriaki Kurita

## Abstract

Negative experiences with medical care have long-term effects on family members’ attitudes and emotions. However, the impact of family members’ experiences on patients’ trust in their physicians and in physicians generally is poorly understood. We aimed to quantify these associations. Our cross-sectional online survey, conducted in Japan during April 2020, involved adults with non-communicable diseases (cardiac disease, diabetes, cancer, depression, and rheumatic disease). The main exposure variable was dissatisfaction with the medical care that family members had received. The main outcomes were patients’ (N=661) trust in their personal physicians and in physicians generally. The study adopted the Japanese version of the Abbreviated Wake Forest Physician Trust Scales. We translated and validated both 5-item scales (general and individual physician trust) for the study. The results showed a lower rating for trust in physicians generally compared to trust in the respondent’s personal physician. Furthermore, dissatisfaction with a family member’s medical care was associated with lower trust in physicians generally. Interestingly, dissatisfaction with a family member’s care was also associated with lower trust in the respondent’s personal physician, but the magnitude of this association was weaker. The lower trust in personal physicians may be mediated by reduced trust in physicians generally. We suggest that physicians enquire about past patients’ negative experiences, including dissatisfaction with family members’ medical care, to repair hidden loss of trust, when they sense that patients doubt them or physicians generally.

## Introduction

Among potentially modifiable patient attitudes, future expectations, or trust, toward physicians (Mechanic, 1998) are important factors in decisions regarding treatment and continued care. Indeed, trust in physicians has been demonstrated to be associated with adherence to medical treatment and continuity of follow-up (Thom et al., 1999). There are two types of trust in physicians: trust in individual physicians and in physicians generally (Hall et al., 2002). Trust in physicians generally can strongly influence the formation of interpersonal physician trust in a specific, known physician, and depends to some extent on an individual’s past experience with their personal physicians (Hall et al., 2002; Rhodes and Strain, 2000).

Previous studies have examined how trust in physicians is affected by many aspects of a patient’s own background or experiences, but very few studies have explored how a family member’s experiences can affect a patient’s trust in physicians. This is a critical gap in our understanding of trust.

Patients can evaluate the quality of family members’ medical care directly through their involvement in their children’s and parents’ medical care (Beernaert et al., 2017; Calabro et al., 2018), especially in intensive or oncological care (Beernaert et al., 2017; Kodali et al., 2014). They can also assess it indirectly through shared medical experiences conveyed by family during everyday communication. Patient and family satisfaction is one of the subjective quality metrics of patient expectations and preferences for medical care experienced by patients and their families (Calabro et al., 2018; Schoenfelder et al., 2011). This is evident from the fact that low satisfaction subsequently influences the health-related behaviors of patients and their families (Schoenfelder et al., 2011) and the possibility of medical litigation claims after unfavorable outcomes (Calabro et al., 2018; Hickson et al., 1994; Stelfox et al., 2005). Therefore, dissatisfaction with family members’ medical care can cause long-term harm to the patient–doctor relationship, resulting not only in behavioral changes in patients but also changes in their attitude toward medical care.

For example, some bereaved children of cancer patients have a long-standing distrust toward the medical care provided to cancer patients (Beernaert et al., 2017). Another study involving family members of patients who had experienced medical errors found that they reported a loss of trust in healthcare and avoidance of medical care in general (Prentice et al., 2020). This system-level loss of trust in healthcare can include a loss of general physician trust (Blendon and Benson, 2001; Hall et al., 2002). Furthermore, general physician trust is said to strongly influence the formation of interpersonal physician trust (Hall et al., 2002). However, trust in an individual physician often stays at a remarkably high level, with patients being willing to forgive physicians they trust (Blendon and Benson, 2001; Hall et al., 2002). However, these possibilities have not been studied extensively. By analyzing the extent to which family members’ dissatisfactory medical experiences influence people’s trust in their own physicians and physicians in general, especially when they themselves are patients, important information can be gained about the hidden origins of distrust.

In Japan, a patient’s family members are usually closely involved in their care. Therefore, it serves as a good setting to investigate the associations between dissatisfaction with family members’ medical care and both general physician trust and interpersonal physician trust.

## Materials and Methods

### Setting and selection

This cross-sectional study was approved by the Ethics Review Board of Kansai University. We used an online panel survey provided by a web-based company (Cross Marketing, Shinjuku-ku, Tokyo) to recruit Japanese participants with non-communicable diseases aged 20 years or older. The respondents were offered a financial incentive. They were discouraged from answering more than once, and researchers could only use their initial response. Figure 1 provides the flow of the study design.

**Figure 1.**
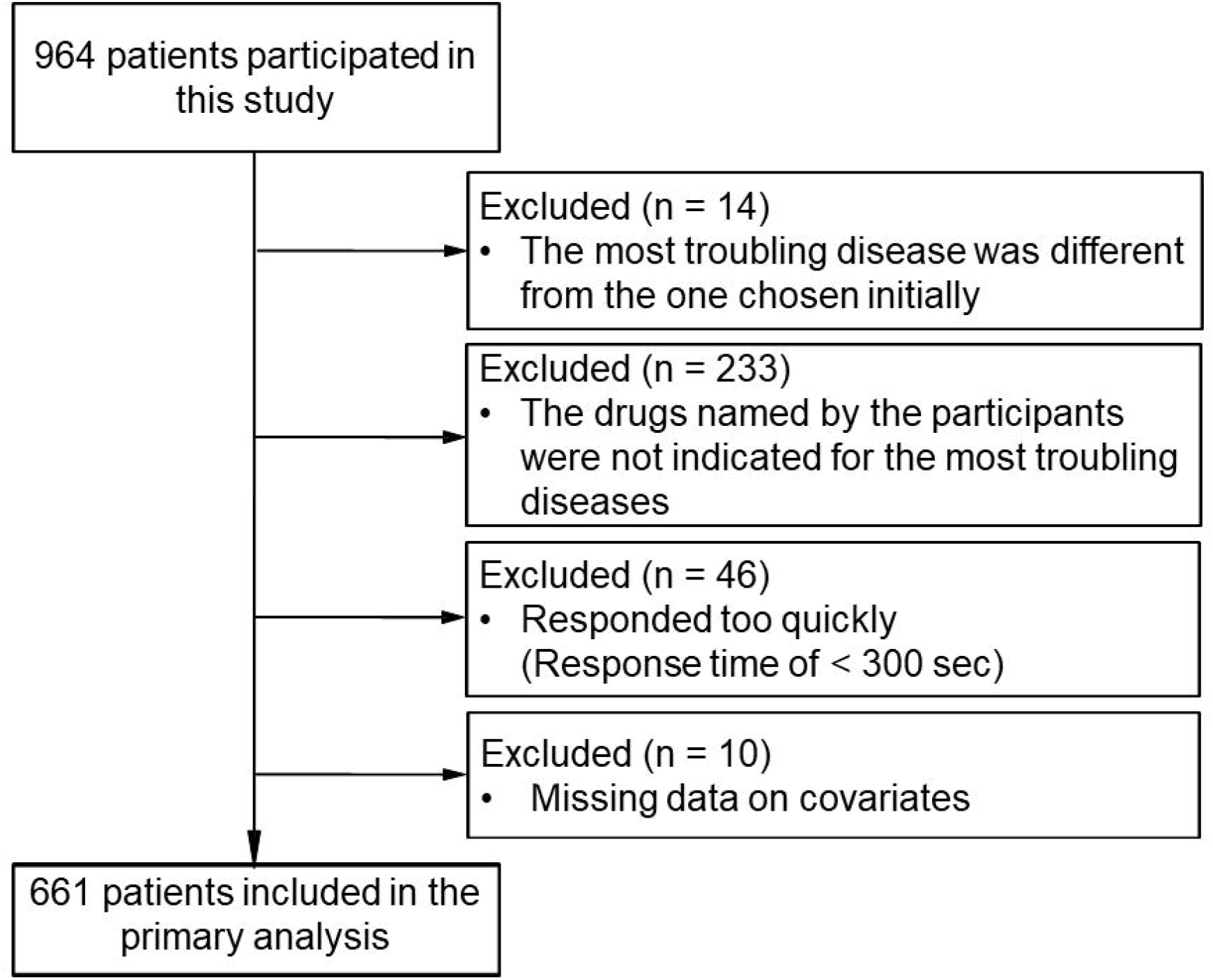
Flow of the study.

### Demographic information

Characteristics such as age, gender, education level, total household income, and zip code were collected using self-reports. We categorized respondents’ prefectures based on the first three digits of the zip code. The duration of the patient–physician relationship was categorized as less than 1 year, 1 to 3 years, and more than 3 years.

### Designing screening items

To prevent random variability and reliability loss through the answers of non-serious respondents, we designed screening items to exclude them from our analysis (Meade and Craig, 2012). As multiple screening items are more effective than a single item, three such items were incorporated before the main survey (Berinsky et al., 2014).

For the first item, respondents had to select a non-communicable disease for which they had received medical treatment twice or more times within the past six months, from a set of eight options. Multiple choices were allowed. For the next item, they had to choose the illness that was most troublesome among those selected in the previous item. If they selected a different disease from the one(s) chosen previously, that meant that either the option(s) chosen from the first item or the second item would have been incorrect; thus, such participants were excluded.

Respondents were then instructed to write the name of a medication prescribed for the most troubling disease in a free-text format. The researchers searched online for label information based on the drug name provided, to assess whether the relevant disease was indicated, in which case the responses were considered valid. Otherwise, the respondents were excluded. However, respondents who chose cancer and wrote “none” for their prescribed drugs were included, as not all cancer treatments require prescribed drugs (e.g., the watch-and-wait method of care is also a reasonable care plan). Two researchers conducted these assessments independently; if the evaluations varied, decisions were reached by consensus.

Respondents were further screened using a response time cut-off since those who responded too quickly may have given spurious answers (Berinsky et al., 2014; Meade and Craig, 2012). Through a pilot test among researchers and their assistants, we found that at least five minutes (300 seconds) were required to complete the survey. Therefore, those respondents taking less than 300 seconds were excluded.

### Dissatisfaction with medical care received by family

Regarding dissatisfaction with the medical care received by their family, the respondents were first given the following instructional statement: “*Looking back on the medical care your family has received, please choose 1 (have had) if you have ever experienced it, or 2 (have not) if you have never experienced it*.” Thereafter, the following question was asked: “*Have you ever been dissatisfied with medical care for a family member’s hospitalization or hospital visit*?”

### Wake Forest Physician Trust Scales: Trust in Doctors Generally and Interpersonal Trust in Physician scales

For this study, short versions of the 5-item “Trust in Doctors Generally” and “Interpersonal Trust in Physician” scales developed by Dugan and Hall (Dugan et al., 2005) were translated into Japanese. The initial translation was performed by two physicians (N.Y. and N.O.), a physician researcher (N.K.), and a quantitative psychologist (T.W.) with experience in scale development. Next, these translations were back translated into English by two bilingual translators (one American and one Canadian) and the wording was compared to the originals to make necessary amendments to the translation. Finally, the back-translated version was sent to the original author (Hall), and additional minor improvements were made. The final versions, approved by the original author, are shown in Supplementary Tables 1 (Interpersonal Trust in a Physician) and 2 (Trust in Physicians Generally).

**Table 1.**
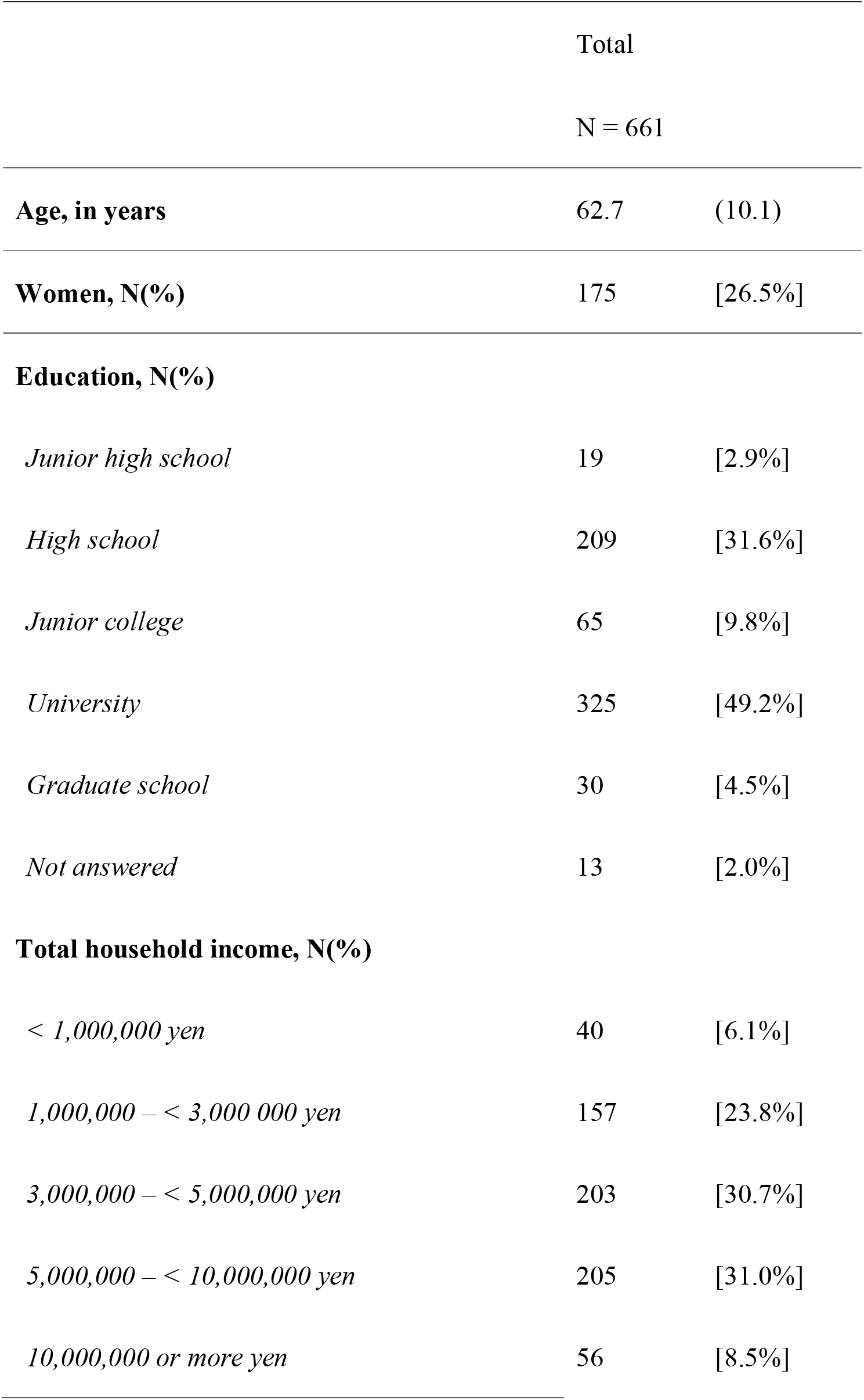
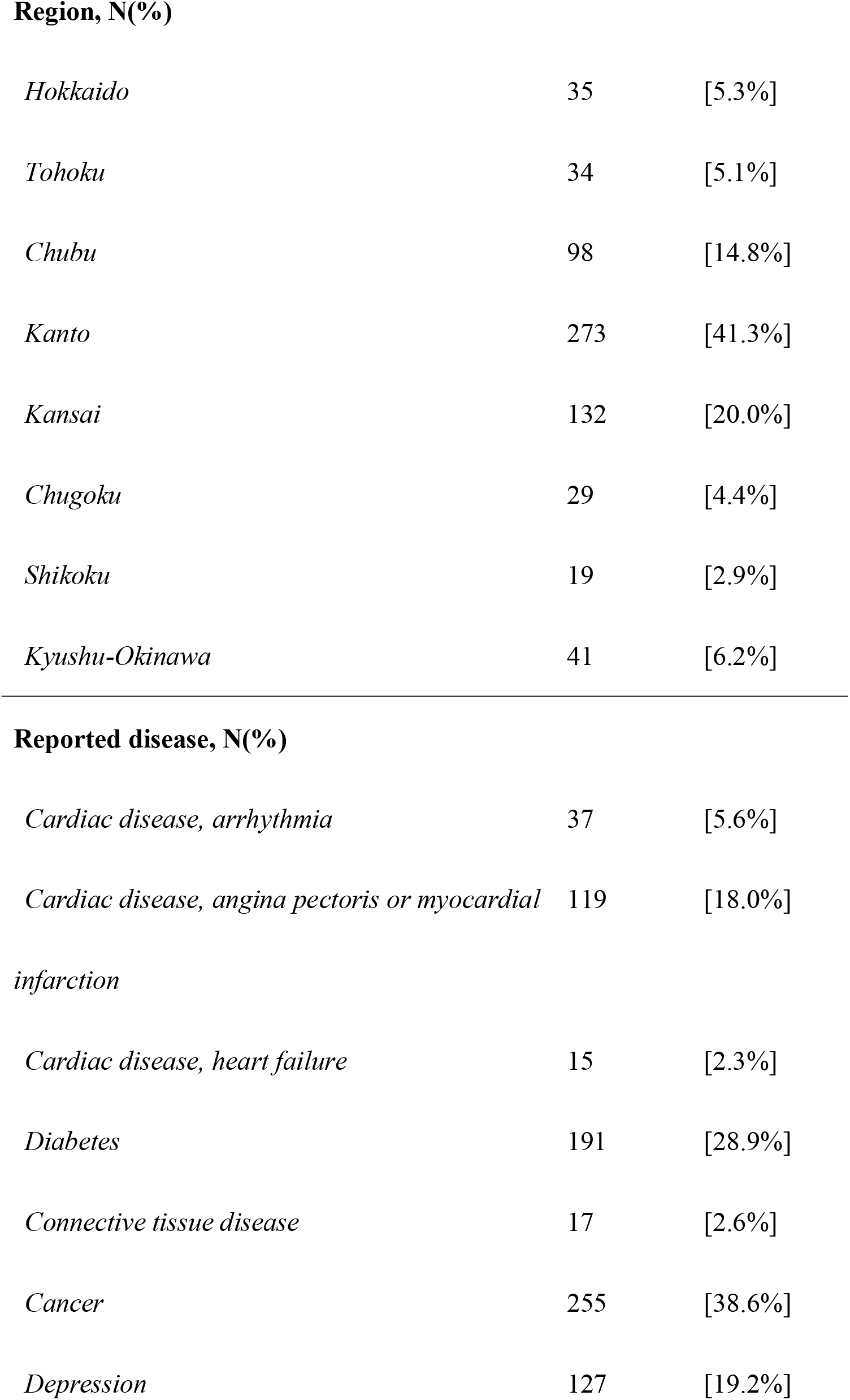
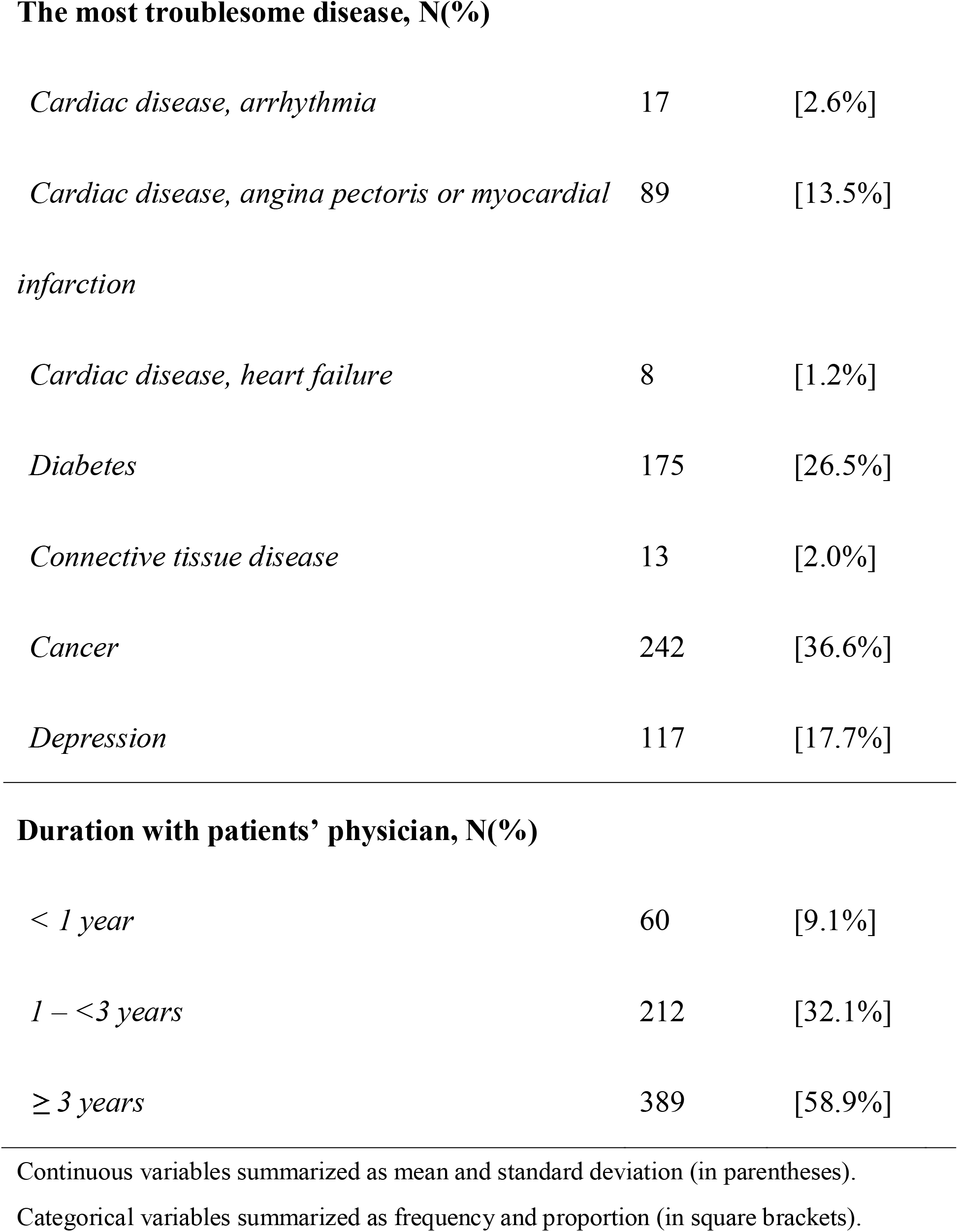
Participant characteristics.

|  | Total |  |
| --- | --- | --- |
|  | N = 661 |  |
| **Age, in years** | 62.7 | (10.1) |
| **Women, N(%)** | 175 | [26.5%] |
| **Education, N(%)** |  |  |
| *Junior high school* | 19 | [2.9%] |
| *High school* | 209 | [31.6%] |
| *Junior college* | 65 | [9.8%] |
| *University* | 325 | [49.2%] |
| *Graduate school* | 30 | [4.5%] |
| *Not answered* | 13 | [2.0%] |
| **Total household income, N(%)** |  |  |
| *< 1,000,000 yen* | 40 | [6.1%] |
| *1,000,000 – < 3,000 000 yen* | 157 | [23.8%] |
| *3,000,000 – < 5,000,000 yen* | 203 | [30.7%] |
| *5,000,000 – < 10,000,000 yen* | 205 | [31.0%] |
| *10,000,000 or more yen* | 56 | [8.5%] |

| **Region, N(%)** |  |  |
| --- | --- | --- |
| *Hokkaido* | 35 | [5.3%] |
| *Tohoku* | 34 | [5.1%] |
| *Chubu* | 98 | [14.8%] |
| *Kanto* | 273 | [41.3%] |
| *Kansai* | 132 | [20.0%] |
| *Chugoku* | 29 | [4.4%] |
| *Shikoku* | 19 | [2.9%] |
| *Kyushu-Okinawa* | 41 | [6.2%] |

| **Reported disease, N(%)** |  |  |
| --- | --- | --- |
| *Cardiac disease, arrhythmia* | 37 | [5.6%] |
| *Cardiac disease, angina pectoris or myocardial infarction* | 119 | [18.0%] |
| *Cardiac disease, heart failure* | 15 | [2.3%] |
| *Diabetes* | 191 | [28.9%] |
| *Connective tissue disease* | 17 | [2.6%] |
| *Cancer* | 255 | [38.6%] |
| *Depression* | 127 | [19.2%] |
| **The most troublesome disease, N(%)** |  |  |
| *Cardiac disease, arrhythmia* | 17 | [2.6%] |
| *Cardiac disease, angina pectoris or myocardial infarction* | 89 | [13.5%] |
| *Cardiac disease, heart failure* | 8 | [1.2%] |
| *Diabetes* | 175 | [26.5%] |
| *Connective tissue disease* | 13 | [2.0%] |
| *Cancer* | 242 | [36.6%] |
| *Depression* | 117 | [17.7%] |
| **Duration with patients' physician, N(%)** |  |  |
| *< 1 year* | 60 | [9.1%] |
| *1 – <3 years* | 212 | [32.1%] |
| *≥ 3 years* | 389 | [58.9%] |

Continuous variables summarized as mean and standard deviation (in parentheses).
Categorical variables summarized as frequency and proportion (in square brackets).

#### Interpersonal Trust in a Physician

Before answering the short version of the Interpersonal Trust in a Physician scale, respondents were instructed as follows: “*Please think of the doctor who cares for your [the most troublesome disease chosen by the participants was automatically displayed here] when you answer these questions. He/she will be considered your doctor for this survey. For the next questions, we are interested in your honest opinion about your doctor. Please choose the answer that best matches your thoughts for each question*.”

#### Trust in Doctors Generally

Before answering the short version of the Trust in Doctors Generally scale, respondents were instructed as follows: “*The following questions may seem similar to the previous ones. However, they are not about your doctor but doctors in general. There is no need to be concerned if you have not thought about these issues before. There is no right or wrong answer. Please choose the answer that best matches your thoughts about doctors in general*.”

For each of the five translated items in each scale, the respondents were instructed to respond on a Likert scale ranging from 1 (strongly disagree) to 5 (strongly agree). We then inverted the score for one negatively-worded item, and changed the sum of the score to a scale ranging from 0 to 100.

### Additional Attitudes

To assess additional attitudes, we used a self-report online questionnaire containing various items. All items were scored on a 5-point Likert scale ranging from 1 (strongly disagree) to 5 (strongly agree).

#### Patient’s general level of interpersonal trust

This was measured using the General Trust Scale (Yamagishi and Yamagishi, 1994). The scale includes six items, with the sum of the items representing the scale score. We expected that both the Interpersonal Trust in a Physician and Trust in Doctors Generally scales would be associated with the General Trust Scale, but the relationships would not be strong, since general interpersonal trust has not previously been found to be strongly related to trust in physicians (Thom et al., 1999).

#### Satisfaction with doctors in general

We assessed this with the item, “*Overall, I am very satisfied with doctors*.”

#### Patients’ satisfaction with their physicians

This was assessed using the item, “*Overall, you are extremely satisfied with your doctor*” (Dugan et al., 2005). Previous studies have strongly correlated it with trust in a physician (Anderson and Dedrick, 1990; Dugan et al., 2005).

#### Patient recommendations about their physicians

This was examined using the item, “*You would recommend your physician to your family and friends*” (Dugan et al., 2005). Higher interpersonal trust scores are considered to be correlated with a better rating of recommendation for their physicians.

#### The desire to change one’s physician

This was examined using the item, “*I have a desire to change my physician*” (Dugan et al., 2005). We expect that the lower the score on interpersonal trust, the stronger will be a patient’s desire to change physicians.

#### Attitude toward adhering to physician treatment

We used one item *(Home treatment is often better than doctor-prescribed medicine* (Freburger et al., 2003)) from a scale that measures skepticism about medical care (Fiscella et al., 1998). We considered that a low level of trust in a physician represents a stronger belief in the effectiveness of home treatment.

### Statistical analysis

All statistical analyses were performed using Stata/SE version 15 (Stata Corp., College Station, TX, USA). Respondents’ characteristics were summarized as means and standard deviations for continuous variables and frequencies and proportions for categorical variables.

For the Interpersonal Trust in a Physician and Trust in Doctors Generally scales, we performed factor analyses with MINRES methods to examine the factorial structures among the 10 combined items. We used the raw scores for reverse items as is. The number of latent factors was assessed by eigenvalues attenuation (Costello and Osborne, 2005). The absolute factor loading magnitudes were calculated. Reliability was assessed by Cronbach’s α and McDonald’s ω coefficients (Dunn et al., 2014). Furthermore, we examined the construct validity of the Interpersonal Trust in Physician scale by testing correlations between the scale and the following factors: patient’s satisfaction with their physician, patient’s recommendations of their physician, general satisfaction with doctors in general, duration of the relationship with their physician (Dugan et al., 2005), and the General Trust Scale. Moreover, we explored the construct validity of the Trust in Doctors Generally scale by testing correlations between it and the Interpersonal Trust in a Physician scale, general satisfaction with doctors, attitude toward adhering to physician treatment, and the General Trust Scale. Spearman’s correlation coefficients were calculated to test these correlations. In addition, to examine whether the respondents rated Interpersonal Trust in a Physician and Trust in Doctors Generally scale items differently, we applied the paired t-test.

Furthermore, to estimate the association between respondents’ dissatisfaction with medical care received by their family and their Trust in Doctors Generally scale score, we fitted a series of linear mixed-effects models with consideration for the clustering effect by prefectures. In unadjusted analysis, only the respondent’s dissatisfaction was fit. In the multivariable-adjusted analysis, the respondent’s dissatisfaction, as well as covariates (age, gender, level of education, total household income, and comorbidities), were fitted to a single model.

We similarly fitted a series of linear mixed-effects models with consideration for the clustering effect by prefectures to examine the respondents’ Interpersonal Trust in a Physician and dissatisfaction with the medical care received by their family. In unadjusted analysis, only the respondent’s dissatisfaction was fit. In the multivariable-adjusted analyses, first, the respondent’s dissatisfaction, as well as covariates (age, gender, level of education, total household income, comorbidities, and duration of relationship between the patient and their physician), were fitted to a single model (adjusted model 1). Second, to assess whether the Trust in Doctors Generally score mediates the relationship between the respondent’s dissatisfaction and Interpersonal Trust in a Physician, covariates in adjusted model 1 plus Trust in Doctors Generally were entered in the linear mixed-effect model (adjusted model 2). These covariates were chosen as they could be associated with both patient dissatisfaction with medical care and trust in physicians.

## Results

The participant characteristics are presented in Table 1. Overall, 964 participants responded, but 303 were excluded, 293 because of the three screener items and 10 because of missing covariates. Subsequently, 661 participants (women: N=175 [26.5%]; Mean age: 62.7±10.1) were included from the primary analysis. The participants’ region of residence extended to 46 prefectures, with Kanto region being the most common (41.3%). The most common troublesome diseases were cancer (36.6%), diabetes (26.5%), depression (17.7%), and heart disease (17.3%).

### Interpersonal Trust in a Physician scale and the Trust in Doctors Generally scale

The eigenvalue attenuation (5.31, 1.37, and 1.05 for the first, second, and third factors, respectively; Supplementary Figure 1) suggested a two-factor solution to the combined 10 items. The absolute values of the factor loadings for items 1 to 5 ranged from 0.44 to 0.86 in factor 1, all of which were above 0.4 (Supplementary Table 3). The absolute values of the factor loadings for items 6 to 10 ranged from 0.42 to 0.89 in factor 2, all of which were above 0.4 (Supplementary Table 3). The inter-factor correlation was moderate (*r* = 0.64). No double loadings between factors occurred for any of the items. Thus, items 1 to 5 could be included in a single factor and reasonably constitute the Japanese version of the Trust in Doctors Generally scale, whereas items 6 to 10 could be included in another single factor and reasonably constitute the Japanese version of the Interpersonal Trust in a Physician scale. Cronbach’s alpha coefficient and McDonald’s omega coefficient were 0.85 and 0.88, respectively, for the Japanese version of the Interpersonal Trust in a Physician scale and 0.88 and 0.93, respectively, for the Japanese version of the Trust in Doctors Generally scale.

For the Interpersonal Trust in a Physician scale (mean: 66.4±17.8), the scores were distributed from 0 to 100, with only 0.2% and 5.3% of them being at the floor and ceiling scores, respectively. As expected, construct validity was supported by the finding that the Interpersonal Trust in a Physician scale was strongly correlated with satisfaction with the physician (ρ = 0.724) and recommending the physician (ρ = 0.678), while it was strongly negatively correlated with the desire to change physicians (ρ = -0.632) (Table 2). Furthermore, this scale was moderately correlated with satisfaction with doctors in general (ρ = 0.550), with a weaker magnitude than that of the correlation between the scale and satisfaction with their physician. The scale was weakly correlated with general interpersonal trust, suggesting that the scale measured a different concept. The scale was not correlated with the duration of the relationship with the physician.

**Table 2.**
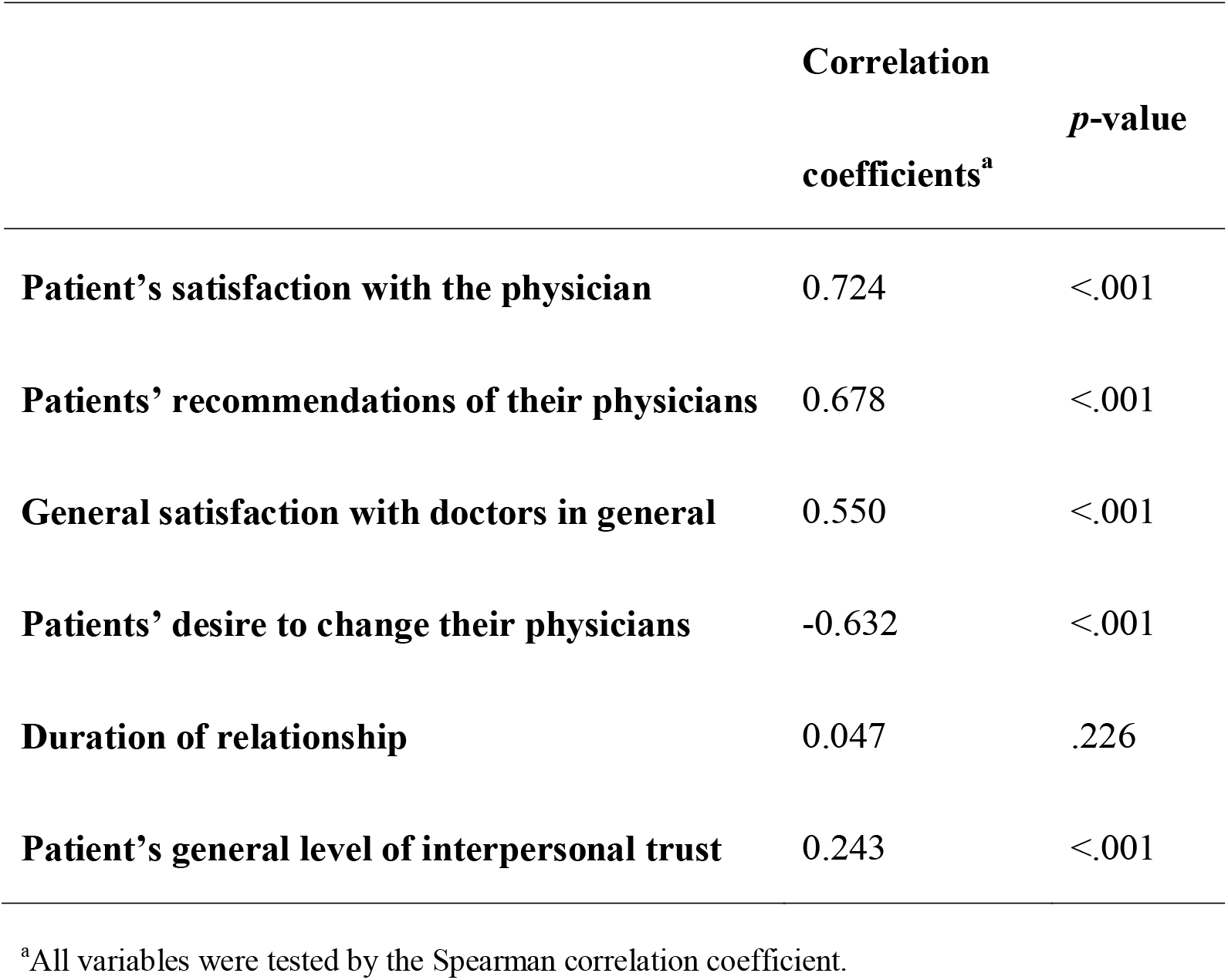
Correlation between interpersonal trust in patient’s physician and selected variables.

|  | **Correlation**<br>**coefficients<sup>a</sup>** | ***p*-value** |
| --- | --- | --- |
| **Patient's satisfaction with the physician** | 0.724 | <.001 |
| **Patients' recommendations of their physicians** | 0.678 | <.001 |
| **General satisfaction with doctors in general** | 0.550 | <.001 |
| **Patients' desire to change their physicians** | -0.632 | <.001 |
| **Duration of relationship** | 0.047 | .226 |
| **Patient's general level of interpersonal trust** | 0.243 | <.001 |

<sup>a</sup>All variables were tested by the Spearman correlation coefficient.

For the Trust in Doctors Generally scale (mean: 57.0±18.4), the scores were distributed from 0 to 100, with only 0.2% and 2.1% of them being at the floor and ceiling scores, respectively. As expected, construct validity was supported by the finding that the Trust in Doctors Generally scale was moderately correlated with satisfaction with doctors in general (ρ = 0.568) (Table 3). The scale was only moderately correlated with the Interpersonal Trust in Physicians (ρ = 0.571) scale, and weakly correlated with general interpersonal trust (ρ = 0.313), suggesting that the scale measured different concepts than these. Furthermore, this scale was weakly negatively correlated with the attitude toward adhering to physician treatment (ρ = -0.213). The scale score was lower than the Interpersonal Trust in a Physician score (*p* < .001).

**Table 3.** Correlation between trust in doctors generally and selected variables.

|  | **Correlation**<br>**coefficients<sup>a</sup>** | ***p*-value** |
| --- | --- | --- |
| **Interpersonal trust in patient's physician** | 0.571 | <.001 |
| **General satisfaction with doctors in general** | 0.568 | <.001 |
| **Believes in home remedies rather than medications**<br>**prescribed by doctors** | -0.213 | <.001 |
| **Patient's general level of interpersonal trust** | 0.313 | <.001 |

<sup>a</sup>All variables were tested by the Spearman correlation coefficient.

### Dissatisfaction with family members’ medical care: relationship with interpersonal trust in a physician and trust in physicians generally

Overall, 233 respondents (35.2%) felt dissatisfaction with family members’ medical care. The association between dissatisfaction with family members’ medical care and trust in doctors generally is shown in Table 4. Dissatisfaction with family members’ medical care was negatively associated with lower trust (mean difference -9.58 [corresponding standardized effect size: -0.52 (Wyrwich et al., 2005)], 95%CI [-12.4 to -6.76]; Figure 2A, adjusted model 1). Older respondents had higher trust scores than did younger respondents (mean difference per 10-year difference: 1.92, 95%CI [0.44 to 3.39]). Those with graduate school education had lower trust compared to those with junior high school education (mean difference: -10.3, 95%CI [-20.4 to -0.17]).

**Table 4.** **Associations between dissatisfaction with medical care provided to patients’ families with Trust in Doctors Generally score**

| Trust in Doctors Generally score,<br>points | Mean<br>difference | (95%CI) |  | p-value |
| --- | --- | --- | --- | --- |
| **<u>Unadjusted model<sup>a</sup></u>** |  |  |  |  |
| **Dissatisfaction with medical care**<br>**provided to a patient’s family** | **-10.4** | **(-13.2** | **to -7.56)** | **<.001** |
| **<u>Multivariable-adjusted model<sup>b</sup></u>** |  |  |  |  |
| **Dissatisfaction with medical care**<br>**provided to a patient’s family** | **-9.58** | **(-12.4** | **to -6.76)** | **<.001** |
| Age, per 10 yr | 1.92 | (0.44 | to 3.39) | .011 |
| Sex, female | -0.81 | (-4.34 | to 2.73) | .654 |
| Education |  |  |  |  |
| *Junior high school* | Ref |  |  |  |
| *High school* | -5.29 | (-13.4 | to 2.85) | .203 |
| *Junior college* | -2.92 | (-12.0 | to 6.12) | .527 |
| *University* | -6.69 | (-14.8 | to 1.41) | .105 |
| ***Graduate school*** | **-10.3** | **(-20.4** | **to -0.17)** | **.046** |
| *Not answered* | -4.94 | (-17.2 | to 7.34) | .430 |
| Total household income |  |  |  |  |
| *< 1,000,000 yen* | -6.16 | (-13.4 | to 1.04) | .094 |
| *1,000,000 – < 3,000,000 yen* | -0.05 | (-5.5 | to 5.42) | .987 |
| *3,000,000 – < 5,000,000 yen* | -0.47 | (-5.7 | to 4.76) | .860 |
| *5,000,000 – < 10,000,000 yen* | -1.56 | (-6.7 | to 3.58) | .552 |
| *10,000,000 or over yen* | Ref |  |  |  |

Analysis of 661 patients in 46 prefectures.
<sup>a</sup>Linear mixed effect models with consideration for prefectural level correlation.
<sup>b</sup>Linear mixed effect model adjusted for age, sex, comorbidities, education, and total household income with consideration for prefectural-level correlation.

**Figure 2.**
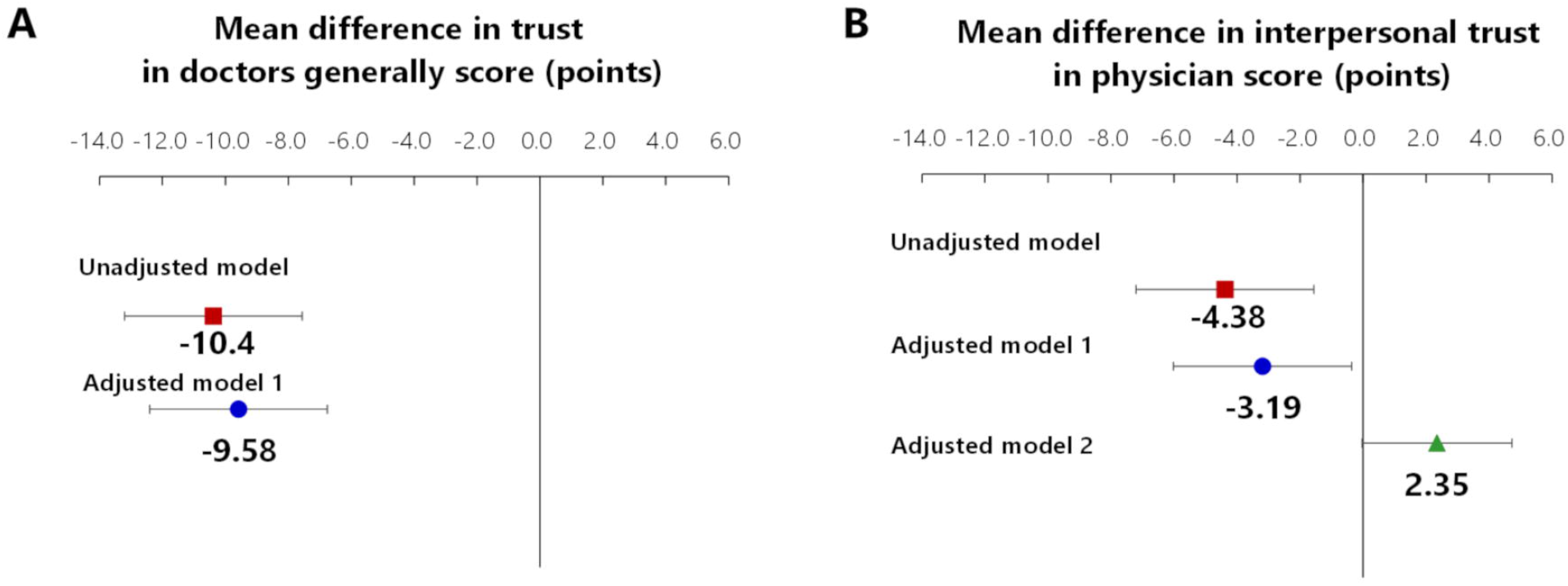
Associations of dissatisfaction with family members’ medical care with the Trust in Doctors Generally score and the Interpersonal Trust in Physician score. Mean differences were estimated using a linear mixed effect model with consideration for prefectural-level correlation using cluster variance. A) Adjusted analysis 1 included age, sex, duration of patient–physician relationship, comorbidities, education, and total household income. B) Adjusted analysis 1 included age, sex, duration of patient–physician relationship, comorbidities, education, and total household income. Adjusted analysis 2 included covariates included in adjusted analysis 1 and Trust in Doctors Generally score. Solid squares, circles, and a triangle indicate point estimates. Error bars indicate 95% CIs.

The association between dissatisfaction with family members’ medical care and interpersonal trust in a physician is shown in Table 5. Similarly to trust in doctors generally, while dissatisfaction with family members’ medical care was negatively associated, the magnitude of the association was very weak (mean difference: -3.19 [corresponding standardized effect size: -0.18 (Wyrwich et al., 2005)], 95%CI [-6.02 to -0.36]; Figure 2B, adjusted model 1). However, the inverse association between dissatisfaction with family members’ medical care and interpersonal trust in a physician disappeared when it was further adjusted by trust in doctors generally (mean difference: 2.35, 95%CI [-0.03 to 4.73]; Figure 2B, adjusted model 2). In this model, respondents who reported general physician trust also had higher trust in their current physicians (mean score difference in Interpersonal Trust in a Physician score per 10-point difference in Trust in Doctors Generally is 5.79 [standardized effect size 0.33; (Wyrwich et al., 2005)], 95%CI [5.17 to 6.42]).

**Table 5.**
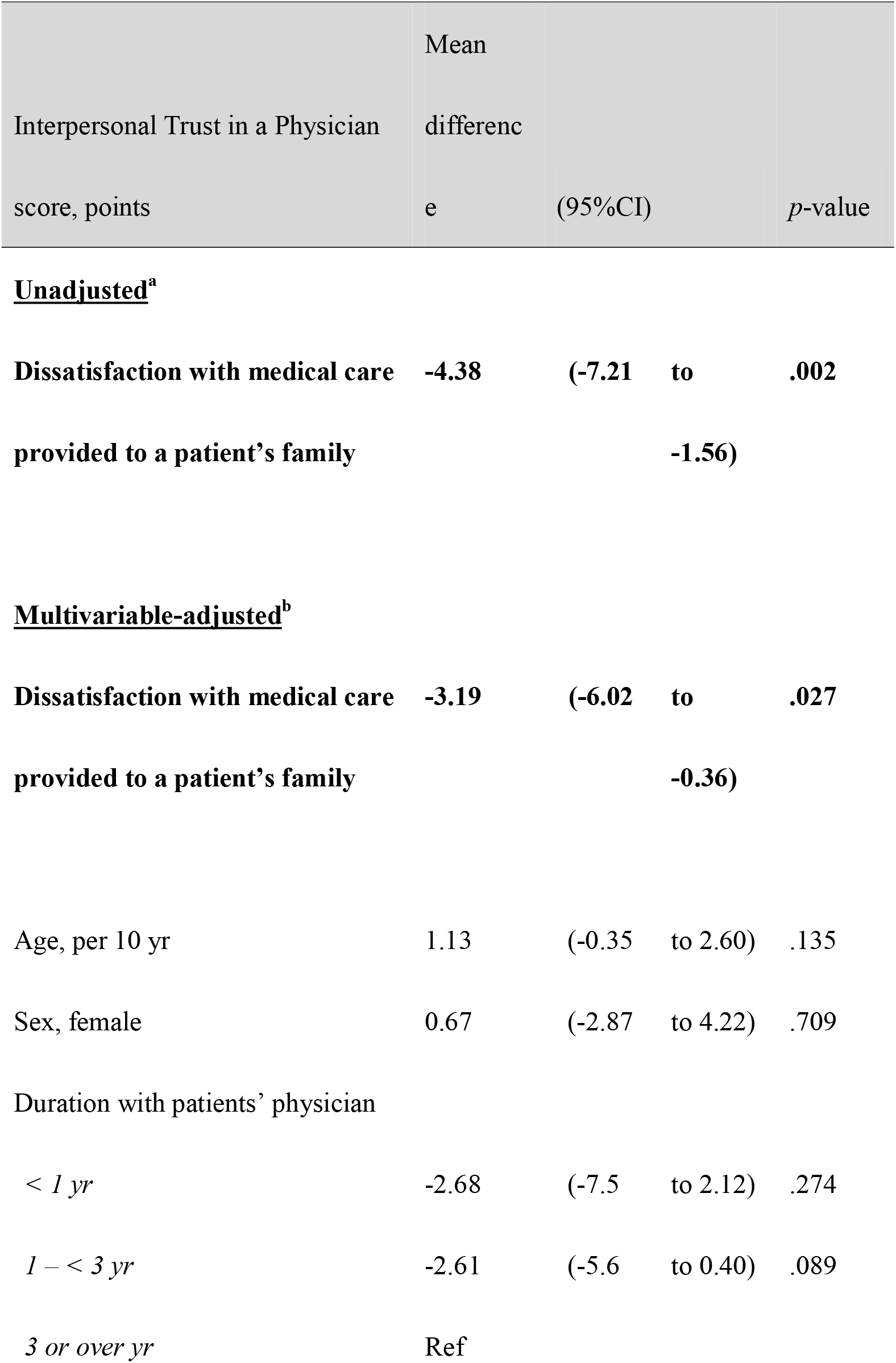
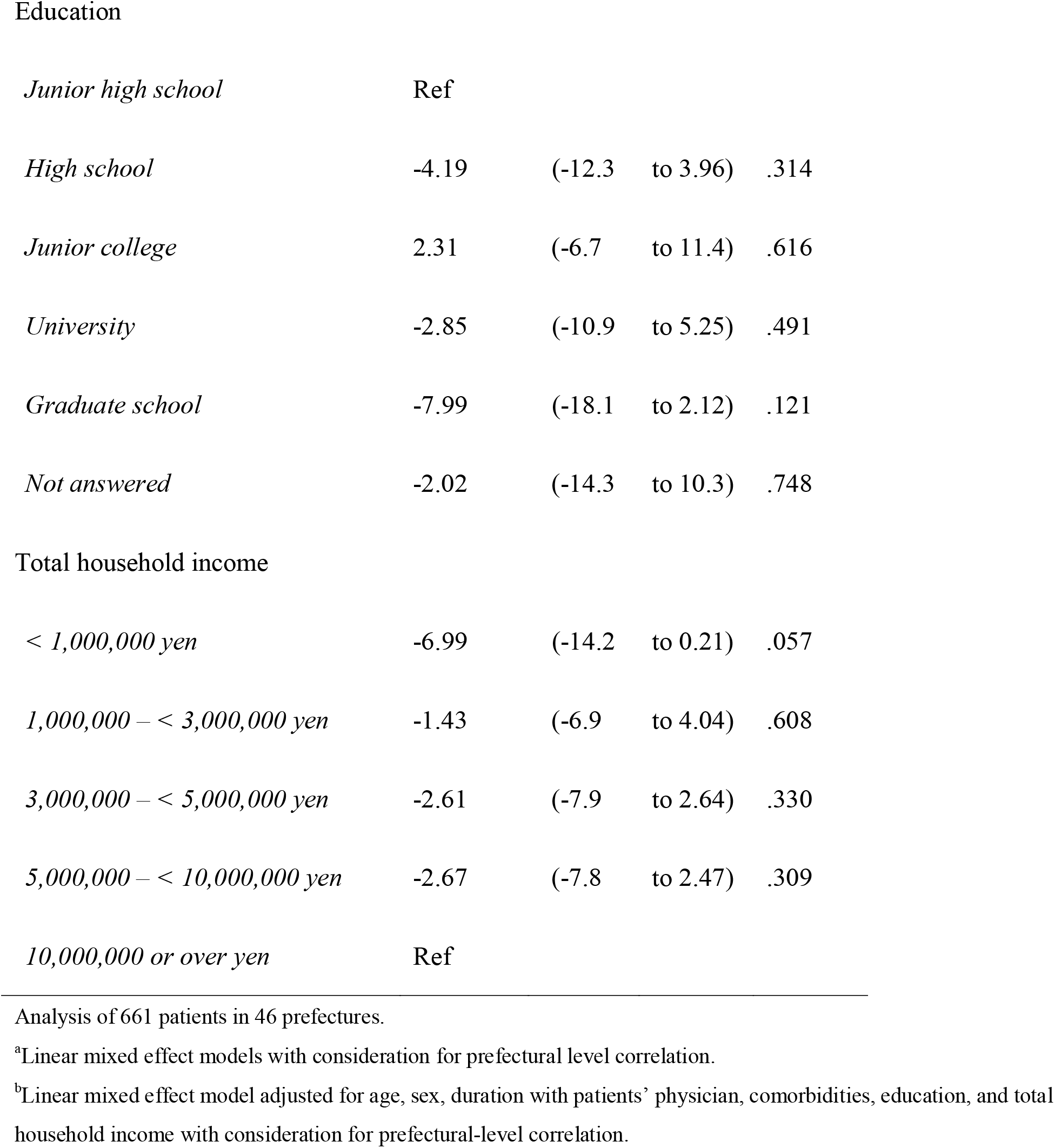
**Associations between dissatisfaction with medical care provided to patients’ families with Interpersonal Trust in a Physician scale**

## Discussion

We examined whether, among patients with non-communicable diseases, dissatisfaction with their family members’ medical care was associated with lower trust in physicians generally, as well as in their own physicians. Past experience of dissatisfaction with family members’ care was associated with a greater reduction in patients’ general physician trust than in trust in their own physicians. In addition, our study suggests that the lower trust in their physicians may be mediated by lower trust in physicians generally. Our findings highlight the importance of researching dissatisfaction with family members’ medical care to identify hidden sources of lost trust in physicians.

In particular, our findings corroborate previous studies and promote insight into trust in physicians. First, a previous study involving the bereaved children of cancer patients revealed long-lasting distrust of the cancer-stricken parents’ medical care among some individuals (Beernaert et al., 2017). However, that study did not examine whether dissatisfaction with the family’s medical care resulting from poor outcomes lowered the children’s trust in their current physicians. Another study that included the family members of patients who had suffered medical errors described a decrease in trust in healthcare at the time, but the study did not quantify the extent to which such negative past experiences affect their current trust in their own physicians and physicians in general (Prentice et al., 2020).

Second, whereas previous reports have indicated that physicians’ image is generally constructed by the media and informal public opinion (Hall et al., 2002; Mechanic, 1998), we were able to show, for the first time, that the individual experience of dissatisfaction with a family member’s medical care is an important factor in reducing general trust in physicians.

Third, our finding that dissatisfaction is associated with a milder decline in trust toward personal physicians than toward physicians in general confirms that interpersonal physician trust is more resilient than trust in the medical profession generally (Blendon and Benson, 2001; Hall et al., 2002).

Last, we found that, among Japanese respondents diagnosed with chronic diseases, interpersonal physician trust was rated higher than trust in physicians in general. This supports the findings of an American study involving a mostly healthy general population (Hall et al., 2002). It indicates that these trust measures are useful across disease types and countries.

Our findings could be useful for physicians and researchers in several ways. First, doctors should consider whether their patients have had any negative medical experiences. This includes dissatisfaction with a family member’s medical care, especially if the patient expresses skepticism toward general medical care or the proposed treatment plan. In doing so, concerns can be addressed. Although the sources of dissatisfaction with family members’ medical care may be broad, including those not attributable to physicians, current physicians can ask about attitudes attributable to past physicians in particular—examples include inquiring about the correctness of a family member’s treatment (Bjertnaes et al., 2012), treatment outcome (Rahmqvist and Bara, 2010), physician’s kindness (Hickson et al., 1994; Schoenfelder et al., 2011), sufficient time with physicians (Hickson et al., 1994; Rahmqvist and Bara, 2010), or participation in decision making (Rahmqvist and Bara, 2010). During these discussions, the doctor should convey compassion, assure patients that not all medical staff are alike, and try not to disappoint them again. After allowing the patient to share the past problem by expressing their anger or anxiety, the doctor should attempt to rebuild a new patient–physician relationship. In particular, active listening and empathy could restore general trust in physicians and strengthen patients’ trust in their current physicians. This was evident in one training program for physicians that focused on communication skills and showed an increase in patients’ satisfaction (Peskin et al., 1995).

Second, we found that the magnitude of a patient’s lower trust associated with past dissatisfaction with family members’ medical care is greater in the case of physicians in general than in that of their own physicians. This may reinforce the pathways of dissatisfaction with family members’ medical care discussed in a previous study (Robinson and Thorne, 1984). Initially, a patient’s family members may naïvely trust that medical professionals would take care of the family member’s illness while understanding the accompanying day-to-day challenges (Robinson and Thorne, 1984). However, the reality of medical care, for example, focusing on the disease rather than the patient as a person, may cause conflict and potential long-term loss of trust in physicians. In addition, trust in a physician a patient knows personally is less likely to be impaired by past negative medical experiences than is trust in physicians generally. This may be attributed to the actions taken by personal physicians to more directly foster interpersonal trust during their practice (Mechanic, 1998).

Third, we found that lower trust in physicians in general may mediate lower trust in current physicians associated with dissatisfaction with family members’ medical care. This supports the notion that trust in physicians generally can influence the formation of interpersonal physician trust (Hall et al., 2002; Rhodes and Strain, 2000).

Our study has several strengths. First, we examined the validity and association among patients with a variety of chronic diseases, including heart disease, diabetes, depression, connective tissue disease, and malignancy. Therefore, our findings about trust in physicians can be applied to a variety of disease settings. Second, by simultaneously conducting a psychometric analysis (i.e., factor analysis) of trust in patients’ physicians and trust in physicians generally, we showed that the concepts of each scale are distinct. Third, we demonstrated for the first time that the mechanism of trust in both individual physicians and physicians in general is similar between the United States and Japan, despite notable differences between these two settings. In Japan, for instance, unlike the United States, all citizens are covered by universal health insurance and have unlimited access to physicians. Thus, our findings support the understanding that both concepts of trust have universal features.

### Limitations

Several limitations of this study warrant a mention. Our study population may not be representative of patients with the same non-communicable diseases because of our survey design. However, we believe that this does not affect the associations between dissatisfaction with family members’ medical care and trust in physicians.

Furthermore, the non-communicable diseases surveyed were based on self-reports and may not have been correctly identified. However, by ascertaining the drug names provided by the respondents and cross-checking them against the chosen diseases, we verified the truthfulness of the diseases that were reported. Another limitation relates to the fact that we did not investigate the reasons for dissatisfaction with family members’ medical care. Thus, the mechanism of lower trust in physicians associated with dissatisfaction could be explored in qualitative studies. Beyond physicians, other factors—including nurses’ care, hospital waiting time, and hygiene—could also influence satisfaction (Bjertnaes et al., 2012). These may be considered in future research.

## Conclusion

In summary, dissatisfaction with family members’ medical care was associated with lower trust among patients in their current physician and physicians generally. The magnitude of lowered trust was greater for physicians in general than for current physicians. Furthermore, the lower trust in current physicians could be mediated by lower trust toward physicians in general. Future research could explore interventions to restore the loss of trust in physicians arising from the dissatisfaction with past medicine experiences, including negative experiences within the family.

## Data Availability

The data that support the findings of this study are available from the corresponding author, [N.K.], upon reasonable request.

## Acknowledgements

We thank Editage (http://www.editage.com) for English language editing.

## Figure Captions

Supplementary Figure 1 Scree plot for the eigenvalues using the response to the combined 10 items of the Interpersonal Trust in a Physician scale and the Trust in Doctors Generally scale

