## Supplemental Table for "Dissatisfaction with family members’ medical care: the relationship with trust in personal physicians and physicians generally in Japan"

Supplementary **Table 1 Japanese version of the Interpersonal Trust in Physician Scale**

Questionnaires in Japanese version are available from the following website ( https://noriaki-kurita.jp/resources/trust-in-physician-jpn/)

| | |
|---|---|
| Instruction sentences | (Original: "If you have several doctors or health professionals that you have seen in the past two years, think of the doctor (or health professional) you know the best when answering the survey. Alternatively, think of the doctor (or health professional) you have seen at least twice in the past two years—we'll just call this person "your doctor." For the next questions, we are interested in your honest opinion about your doctor. For each of these questions, please tell me whether you strongly agree, agree, are neutral, disagree, or strongly disagree.") |
| Question 1 | (Original: "Sometimes, your doctor cares more about what is convenient for (him/her) than about your medical needs.") |
| Question 2 | (Original: "Your doctor is extremely thorough and careful.") |
| Question 3 | (Original: "You completely trust your doctor's decisions about which medical treatments are best for you.") |
| Question 4 | (Original: "Your doctor is totally honest in telling you about all of the different treatment options available for your condition.") |
| Question 5 | (Original: "All in all, you have complete trust in your doctor.") |
| Response options for Questions | (Original: strongly disagree/disagree/neutral/agree/strongly agree) |

The original English version (Dugan et al., 2005) is also provided for each item and response. Before using this instrument, please register through https://***.jp/resources/trust-in-physician-jpn/. In addition, please cite this article as a reference:

2021. Dissatisfaction with family members' medical care: the relationship with trust in personal physicians and physicians generally in Japan. Soc Sci Med.

*The instructional statements generated through the formal process of translation are presented. Modified instructional statements were used in the online survey of this study as follows: “*Please answer this section of the survey while thinking about the doctor who is taking care of your [the most troublesome disease chosen by the participants was automatically displayed in this place]. We will refer to him/her as your doctor. The following questions concern your feelings about your doctor. Please choose the answer that best matches your thoughts for each question.*”

**Supplementary Table 2 Japanese version of the Trust in Doctors Generally Scale**

Questionnaires in Japanese version are available from the following website ( https://noriaki-kurita.jp/resources/trust-in-physician-jpn/)

| | |
|---|---|
| Instruction sentences | (Original: "These next questions may sound similar to the ones I asked you before. However, these are about DOCTORS IN GENERAL, not your doctor. You may not have thought about these issues before, but do not worry; there are no right or wrong answers. How much do you agree or disagree with the following statements about DOCTORS IN GENERAL?") |
| Question 1 | (Original: "Sometimes doctors care more about what is convenient for them than about their patients' medical needs.") |
| Question 2 | (Original: "Doctors are extremely thorough and careful.") |
| Question 3 | (Original: "You completely trust doctors' decisions about which medical treatments are best.") |
| Question 4 | (Original: "A doctor would never mislead you about anything.") |
| Question 5 | (Original: "All in all, you trust doctors completely.") |
| Response options for Question | (Original: strongly disagree/disagree/neutral/agree/strongly agree) |

The original English version (Dugan et al., 2005) is also provided for each item and response. Before using this instrument, please register through https://***.jp/resources/trust-in-physician-jpn/. In addition, please cite this article as a reference:

2021. Dissatisfaction with family members' medical care: the relationship with trust in personal physicians and physicians generally in Japan. Soc Sci Med.

*The instructional statements generated through the formal process of translation are presented. Modified instructional statements were used in the online survey of this study as follows: "*The following questions may seem similar to the previous ones. However, they are not about your doctor but doctors in general. There is no need to be concerned if you have not thought about these issues before. There is no right or wrong answer. Please choose the answer that best matches your thoughts about doctors in general.*"

**Supplementary Table 3 Descriptive statistics and factor loadings of the combined 10 items of the Interpersonal Trust in a Physician scale and the Trust in Doctors Generally scale**

| Item No. | Origin | Questions | Mean | SD | Factor loading | |
|---|---|---|---|---|---|---|
| | | | | | Factor 1 | Factor 2 |
| 1 | g4 | A doctor would never mislead you about anything. | 3.25 | 0.91 | 0.86 | -0.11 |
| 2 | g3 | You completely trust doctors' decisions about which medical treatments are best. | 3.32 | 0.89 | 0.86 | 0.05 |
| 3 | g5 | All in all, you trust doctors completely. | 3.43 | 0.88 | 0.85 | 0.03 |
| 4 | g2 | Doctors are extremely thorough and careful. | 3.18 | 0.89 | 0.82 | 0.01 |
| 5 | g1 | Sometimes doctors care more about what is convenient for them than about their patients' medical needs. | 2.78 | 0.92 | -0.44 | -0.08 |
| 6 | i5 | All in all, you have complete trust in your doctor. | 3.74 | 0.92 | -0.03 | 0.89 |
| 7 | i2 | Your doctor is extremely thorough and careful. | 3.47 | 0.89 | 0.08 | 0.78 |
| 8 | i3 | You completely trust your doctor's decisions about which medical treatments are best for you. | 3.68 | 0.88 | 0.11 | 0.76 |

| | | | | | | |
|---|---|---|---|---|---|---|
| 9 | i4 | Your doctor is totally honest in telling you about all of the different treatment options available for your condition. | 3.75 | 0.85 | 0 | 0.74 |
| 10 | i1 | Sometimes, your doctor cares more about what is convenient for (him/her) than about your medical needs.。 | 2.36 | 0.98 | 0.03 | -0.42 |

The absolute values of factor loadings for items no. 1 to no. 5 ranged from 0.44 to 0.86 in factor 1, all of which were above 0.4. The absolute values of factor loadings for items no. 6 to no. 10 ranged from 0.42 to 0.89 in factor 2, all of which were above 0.4. The inter-factor correlation was moderate (r = 0.64). In each item, no double loadings between factor 1 and factor 2 occurred. Thus, items no. 1 to no. 5 could be included in a single factor and reasonably constitute the Japanese version of the Trust in Doctors Generally scale, whereas items no. 6 to no. 10 could be included in another single factor and reasonably constitute the Japanese version of the Interpersonal Trust in a Physician scale.

**Supplementary Figure 1 Scree plot for the eigenvalues using the response to the combined 10 items of the Interpersonal Trust in a Physician scale and the Trust in Doctors Generally scale**

The eigenvalue attenuation was largest between the first and second factors (5.31, 1.37, and 1.05 for the first, second, and third factors, respectively), indicating that the items had two dimensions.

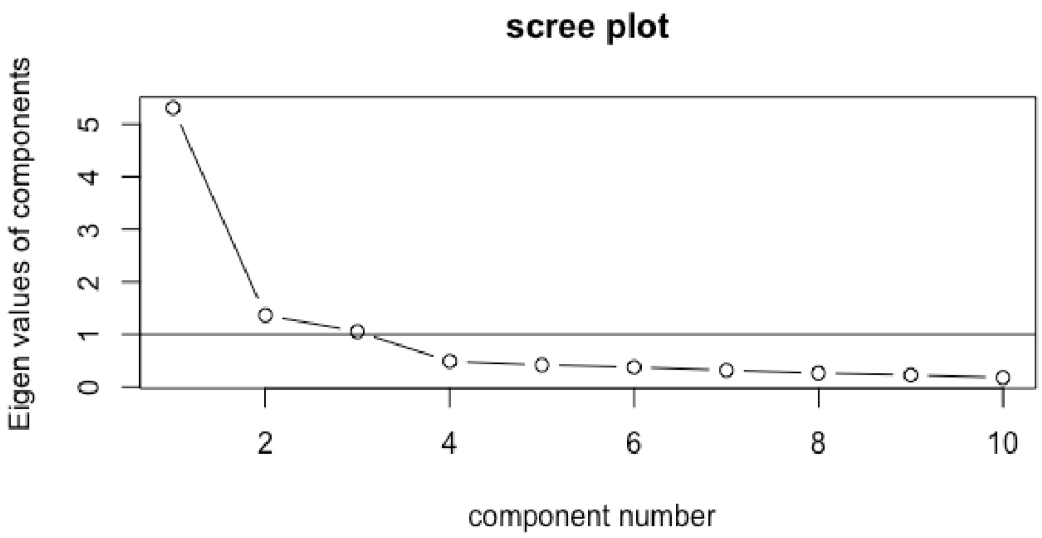
